# Atrial functional substrates for the prediction of atrial fibrillation recurrence after pulmonary vein isolation

**DOI:** 10.1101/2023.10.13.23297031

**Authors:** Masaharu Masuda, Yasuhiro Matsuda, Hiroyuki Uematsu, Mitsutoshi Asai, Shin Okamoto, Takayuki Ishihara, Kiyonori Nanto, Takuya Tsujimura, Yosuke Hata, Naoko Higashino, Sho Nakao, Toshiaki Mano

## Abstract

**Background:** Low-voltage areas have been used as atrial structural substrates in estimating fibrotic degeneration in patients with atrial fibrillation (AF). The high-resolution maps obtained by recently developed mapping catheters allow the visualization of several functional abnormalities.

**Objectives:** We investigated the association between left atrial (LA) functional abnormal findings on a high-resolution substrate map and AF recurrence in patients who underwent pulmonary vein isolation without any additional LA substrate ablation.

**Methods:** One hundred consecutive patients who underwent second ablation for AF (paroxysmal, 48%; persistent, 52%) were considered for enrollment. Patients with extra-pulmonary-vein LA substrate ablation during the initial and second ablation were excluded. LA mapping was performed using a 64-pole mini-basket catheter on the Rhythmia mapping system. Patients were followed for 2 years.

**Results:** AF recurrence developed in 39 (39%) patients. On the high-resolution substrate map, AF recurrence was associated with the presence of the following findings: low-voltage areas (<1.0 mV, >5cm^2^; hazard ratio [HR]=2.53; 95% confidence interval [CI]=1.30-4.93; p<0.006), fractionated-electrogram areas (≥5 peaks, >5cm^2^; HR=2.15, 95%CI=1.10-4.19; p=0.025), LA conduction time of >130 msec (HR=3.11, 95%CI=1.65-5.88, p<0.0001), deceleration zone (≥5 isochrones/cm^2^; HR=1.97, 95%CI=1.04-3.37, p=0.039), and multiple septal breakout points (HR=3.27, 95%CI=1.50-7.16, p=0.003). Accumulation of these risk factors increased AF recurrence in a stepwise manner, with an HR=1.90, 95%CI=1.44-2.52, p<0.00001 for each additional risk factor.

**Conclusion:** A high-resolution map revealed new LA functional substrates associated with AF recurrence. Implementation of functional substrates may improve the prediction of AF recurrence after ablation, and possibly aid the development of tailored AF ablation strategies.

**Graphical abstract:** Abnormal LA substrates and accumulation of risk factors
Study design, representative maps demonstrating abnormal substrates, and AF recurrence rates stratified by the number of risk factors are shown. Accumulation of these risk factors increased AF recurrence rates in a stepwise manner with an HR = 1.90, 95% CI = 1.44 - 2.52, p<0.00001, for 1 risk increase. AF indicates atrial fibrillation; HR, hazard ratio; CI, confidence interval.

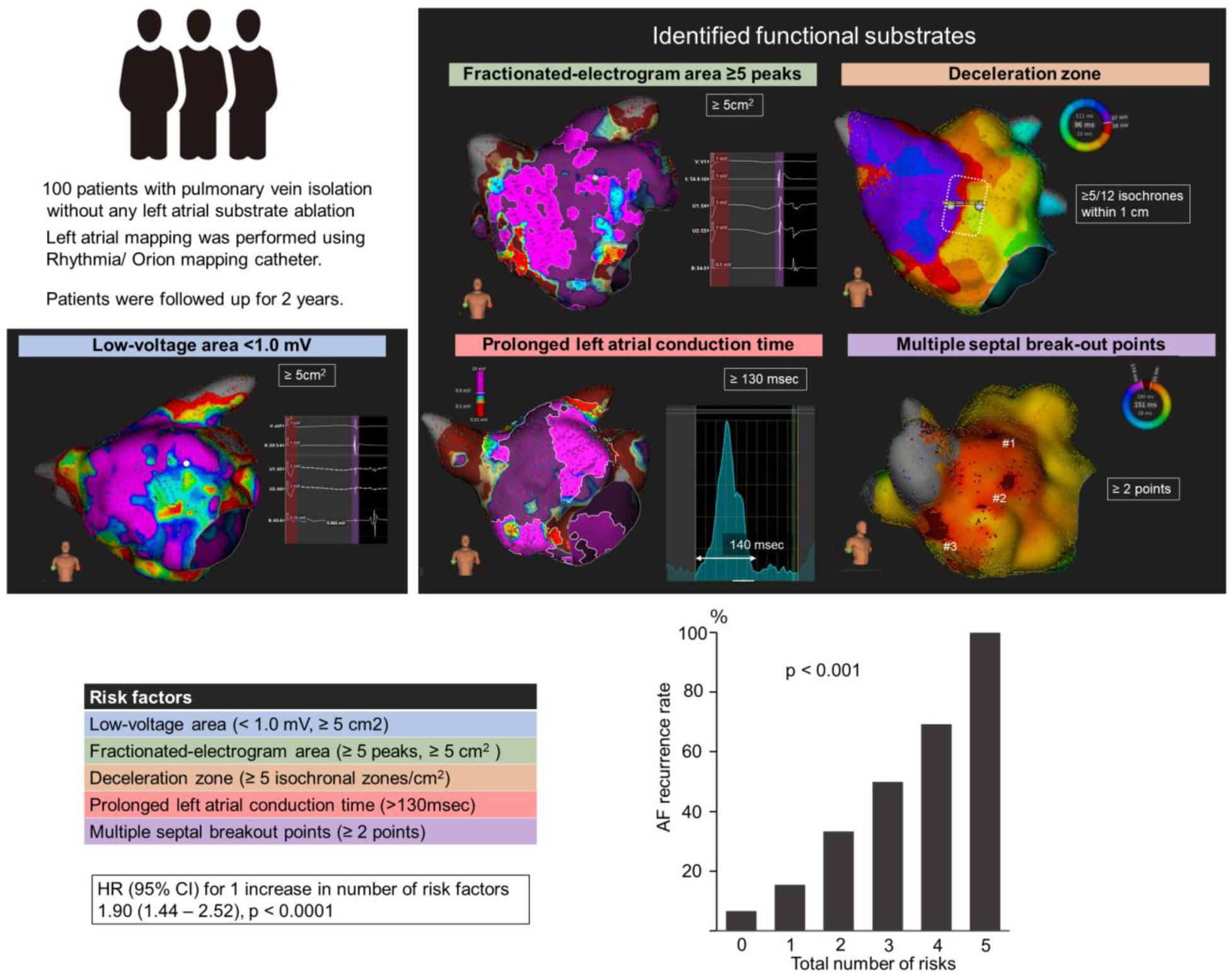

## Introduction

Pulmonary vein isolation is a cornerstone of atrial fibrillation (AF) ablation.^1^ However, a considerable number of patients experience recurrence of AF. The main causes of AF recurrence following initial AF ablation are advanced atrial remodeling in addition to electrical reconnection between the left atrium to pulmonary vein.

Identifying patients who need additional left atrial (LA) ablation after pulmonary vein isolation is critically important. In this respect, the presence of left atrial (LA) low-voltage areas on a substrate map has been demonstrated to correlate well with atrial myocardial degeneration, including fibrosis,^2^ and is a predictor of recurrence of atrial tachyarrhythmias after pulmonary vein isolation.^3–5^ However, atrial bipolar voltage is not enough to understand the atrial arrhythmogenic substrate, or to predict rhythm outcome with sufficient accuracy.

Recent innovations in 3-dimentional mapping systems and mapping catheters have increased mapping density and recorded signal quality, resulting in high-resolution substrate maps which enable the depiction of more detailed substrate, such as local electrogram fractionation and abnormal propagation pattern. These functional substrates may represent atrial electrical remodeling which predisposes to AF development, and in turn enable the prediction of AF recurrence after pulmonary vein isolation.

Here, we investigated the association between LA functional substrates and long-term rhythm outcomes in patients who underwent pulmonary vein isolation.

## Methods

### Patients

From September 2016 to August 2019, consecutive patients who underwent a second ablation for AF with LA mapping using the RHYTHMIA system (Boston Scientific, Marlborough [Cambridge] MA, USA) at our institution were considered for enrollment. To avoid the influence of adjunctive substrate ablation in addition to pulmonary vein isolation, patients who underwent extra-pulmonary-vein ablation at the initial or second ablation were excluded (Supplementary Figure 1). Other exclusion criteria were inappropriate substrate mapping, age < 20 years, and prior cardiac surgery.

This study complied with the Declaration of Helsinki. The protocol was approved by our institutional review board, and written informed consent for the ablation and participation in the study was obtained from all patients.

### Substrate mapping

Electrophysiological studies and catheter ablation were performed under general anesthesia with propofol and fentanyl. A 6-Fr decapolar electrode was inserted into the coronary sinus and a second 6-Fr decapolar electrode was placed in the right atrium. Following transseptal puncture at the fossa ovalis, two long sheaths were introduced into the left atrium using a single transseptal puncture technique. Mapping and ablation were then performed under guidance from the electroanatomical mapping system (RHYTHMIA*^®^*).

Prior to ablation, substrate mapping was performed under paced rhythm from the high right atrium or great cardiac vein. A mini-basket array catheter with 64 mini-electrodes (IntellaMap Orion^®^, Boston Scientific) was moved to draw the whole LA surface. Mapping was continued until coverage of the entire LA surface using an interpolation threshold of 5 mm. Adequate endocardial contact was confirmed by stable electrograms and distance to the geometry surface. The criteria used for beat acceptance included stable cycle length, stable timing difference between two reference electrodes placed in the coronary sinus, respiratory gating, stable catheter location, and a stable catheter signal compared to adjacent points. The confidence mask and projection distance were set at 0.03 mV and 2 mm, respectively, and band pass filter was set at 30 to 500 Hz.

### Catheter ablation

Following substrate mapping, ablation was performed. When electrical reconnection between left atrium and pulmonary vein was observed, re-isolation of pulmonary vein was performed by radiofrequency application at the conduction gaps. Atrial burst pacing (cycle length 200 to 300 ppm for 5 sec) was performed, followed by a high-dose isoproterenol provocation test (infusion of 5, 10, and 20 µg/min isoproterenol for 2 min each) to induce AF or atrial tachycardia. AF-triggering ectopies or frequent ectopies originating from the superior vena cava (SVC) were eliminated by circumferential SVC isolation. Non-PV/SVC AF-triggering ectopies were ablated at the earliest activation site. A spontaneously developed or induced AT was also mapped using the electroanatomical mapping system. Focal ablation targeting the earliest activation site for a centrifugal AT or ablation crossing the reentrant circuit for a macro-reentrant AT was performed. As mentioned above, patients with LA ablation were excluded from enrollment.

### Assessment of LA substrate

Any of the following abnormal findings were measured on an offline work station by a medical engineer blinded to rhythm outcomes. Isolated areas encircled by ablation lesions for pulmonary vein isolation were not included in measurement. Representative maps demonstrating abnormal substrates are shown in the Figure 1.

**Figure 1.**
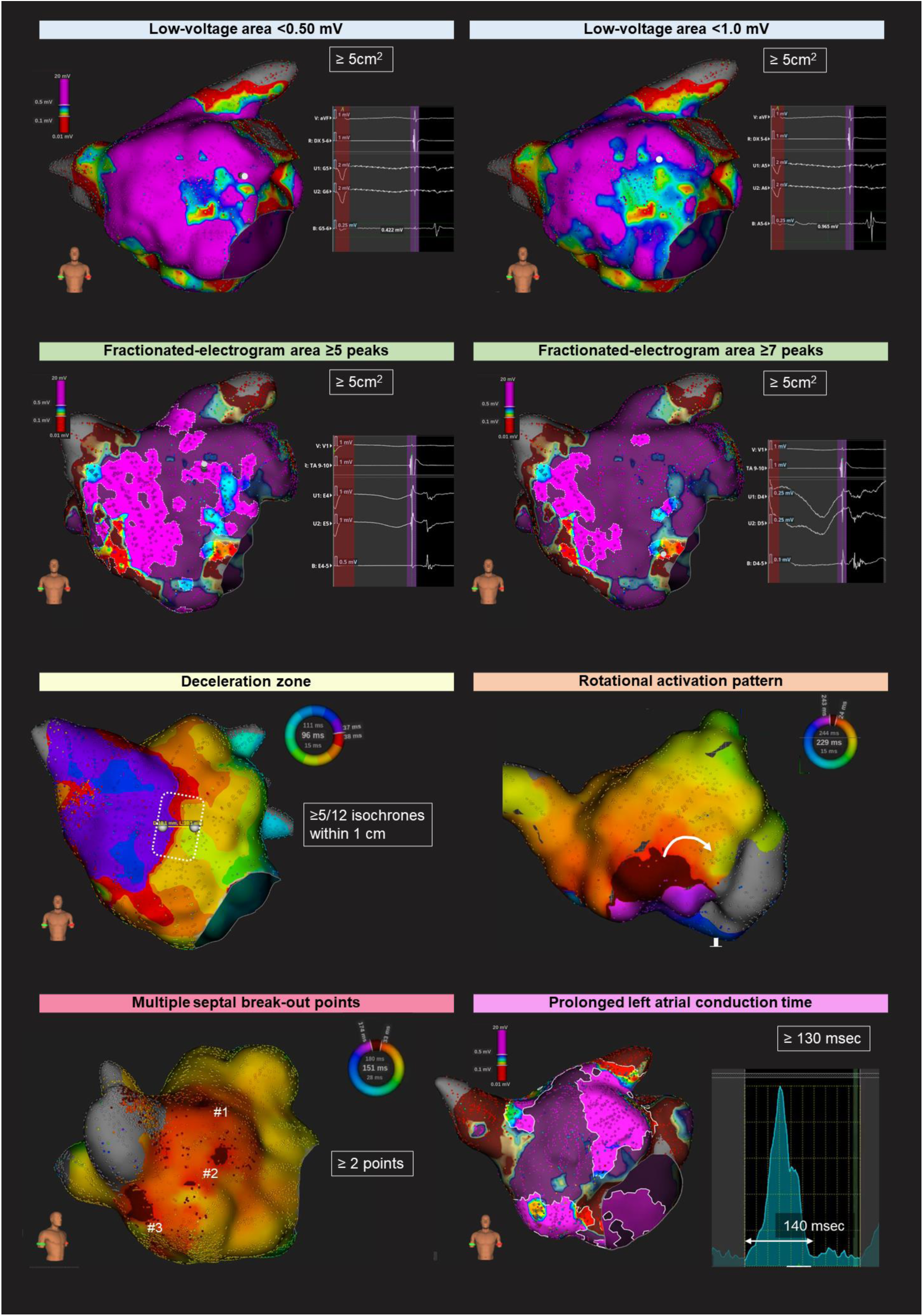
Abnormal LA substrates. Representative maps demonstrating abnormal substrates are shown. Low-voltage areas were defined as areas with a bipolar voltage < 0.50 mV or 1.0 mV. Fractionated-electrogram areas were defined as areas where fractionated signals had ≥ 5 or 7 peaks. Deceleration zones were defined as regions with isochronal crowding of ≥ 5 isochrones within a 1-cm radius among 12 isochrones which evenly divide the entire LA activation time. Rotational activation patterns were defined as sites showing wavefront propagation rotating > 90° (inward curvature) directly above or at the edge of the isochronal crowding zone. Multiple septal break-out points were defined when the number of left septal activation break-out points was ≥ 2. An LA conduction time of ≥ 130 msec was considered prolonged.

Low-voltage areas were defined as areas with bipolar voltage < 0.50mV or 1.0 mV, and were highlighted by setting the threshold values of the color bar on voltage map. The size of the low-voltage areas was measured by manually tracing the border of the areas.

Fractionated-electrogram areas were defined as areas where fractionated signals were recorded, and were highlighted under complex activation mode in the LUMIPOINT module. The size of fractionated areas was measured by manually tracing the border of the areas. The number of peaks used to determine a fractionated signal was set at 5 or 7.

To assess local conduction speed, the activation map was displayed with 12 equally distributed isochrones covering the entire LA activation time. Deceleration zones were defined as regions with isochronal crowding of ≥ 4 or ≥ 5 isochrones within a 1-cm radius. Rotational activation pattern was defined in sites showing wavefront propagation rotating > 90° (inward curvature) directly above or at the edge of the isochronal crowding zone.

Septal propagation was assessed by counting the number of break-out points on the left septal wall. This measurement was performed only in patients with pacing from the right atrium. Wavefront propagation was assessed by activation search mode in the LUMIPOINT module. Multiple septal break-out points were recognized when the number of break-out points was ≥ 2 or 3. Supplementary video 1 demonstrates a propagation map with multiple septal break-out points and rotational activation pattern.

LA conduction time was calculated as the time difference between the start of LA activation, usually at the LA septum, and the end of LA activation, usually at the lateral mitral annulus. Prolongation of LA conduction time was defined when this value was ≥ 130 msec.

### Follow-up

Patients were followed up every 2 to 4 weeks at the dedicated arrhythmia clinic of our institution. Routine ECGs were obtained at each outpatient visit, and 24-h ambulatory Holter monitoring was performed every 3 months post-ablation. When patients experienced symptoms suggestive of an arrhythmia, a surface ECG, ambulatory ECG, and/or cardiac event recording were also obtained. Any of the following events from the initial 3 months after the ablation (blanking period) were considered to indicate AF recurrence: (1) atrial tachyarrhythmias recorded on a routine or symptom-triggered ECG during an outpatient visit; or (2) atrial tachyarrhythmias of at least 30-sec duration on ambulatory ECG monitoring. No antiarrhythmic drugs were prescribed 3 months post-ablation unless recurrent atrial tachyarrhythmias were observed.

### Statistical analysis

Continuous data are expressed as the mean ± standard deviation (SD) or median (interquartile range). Categorical data are presented as absolute values and percentages. Tests for significance were conducted using the unpaired t-test or nonparametric test (Mann-Whiney U test) for continuous variables, and the chi-square test or Fisher’s exact test for categorical variables. Cox proportional hazard models were used to determine parameters on the substrate map that were associated with AF recurrence. To estimate the optimal cut-off values for the prediction of AF recurrence, receiver-operating characteristic curves were constructed for low-voltage areas, fractionated-electrogram area sizes and LA conduction time. Survival rates free from atrial tachyarrhythmias were calculated using the Kaplan-Meier method. Comparison of survival curves between groups was assessed with the 2-sided Mantel-Haenszel (log-rank) test. All analyses were performed using SPSS version 15.0 software (SPSS, Inc., Chicago, IL, USA).

## Results

### Baseline and procedural characteristics

During the study period, 270 patients underwent second ablation for AF using the Rhythmia system (Supplementary Figure 1). Among these, 156 patients with LA ablation outside of pulmonary veins and 14 with an inappropriate substrate map were excluded. Finally, 100 patients were enrolled in the study. Mapping was performed under pacing from the right atrium (81%) and great cardiac vein (19%).

During 2-year follow-up, 39 (39%) patients developed AF recurrence. Baseline characteristics were compared between those with and without AF recurrence (Table 1). Patients with AF recurrence were more likely to have long-standing persistent AF and a history of stroke or thromboembolism; and had a higher CHA_2_DS_2_VASc score, N-terminal prohormone of brain natriuretic peptide level, and larger left atrium.

**Table 1.**
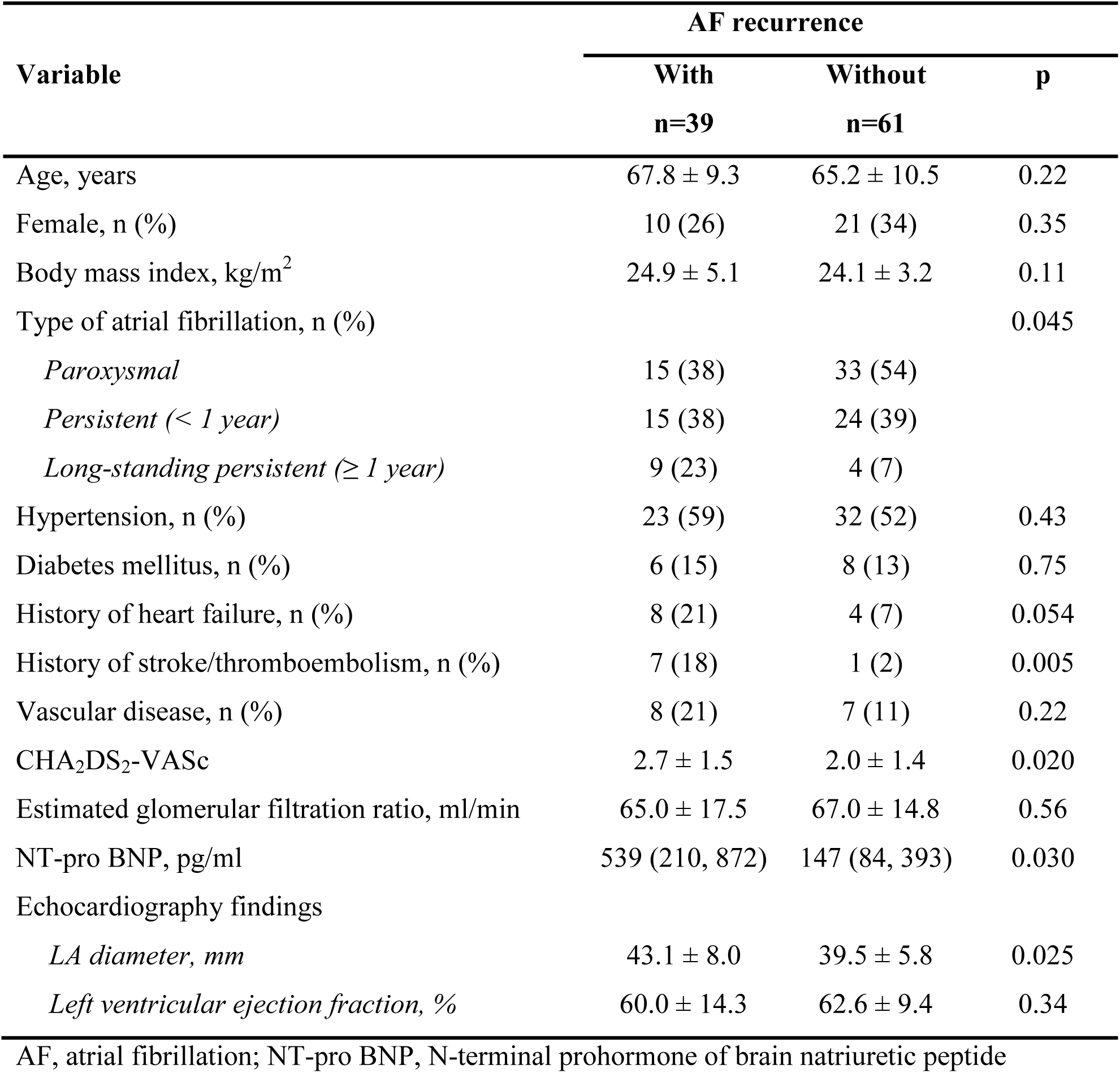
Patient characteristics.

**Table 2.** Ablation procedures.

| Variable | AF recurrence |  | p |
| --- | --- | --- | --- |
|  | With<br>n=39 | Without<br>n=61 |  |
| Initial ablation |  |  |  |
| Energy source for pulmonary vein isolation, n (%) |  |  | 0.039 |
| *Radiofrequency energy* | 33 (85) | 40 (66) |  |
| *Cryo energy* | 6 (15) | 21 (34) |  |
| Achievement of pulmonary vein isolation, n (%) | 39 (100) | 61 (100) | 1.0 |
| Cavotricuspid isthmus ablation, n (%) | 8 (21) | 10 (16) | 0.60 |
| Isolation of superior vena cava, n (%) | 1 (3) | 2 (3) | 1.00 |
| Non-pulmonary-vein trigger ablation, n (%) | 2 (5) | 4 (7) | 1.00 |
| Second ablation |  |  |  |
| Mapping points, points | 6,431 ± 2,377 | 5,363 ± 1,792 | 0.012 |
| Re-isolation of reconnected pulmonary vein, n (%) | 26 (67) | 47 (77) | 0.25 |
| Cavotricuspid isthmus ablation, n (%) | 8 (21) | 16 (26) | 0.11 |
| Isolation of superior vena cava, n (%) | 10 (26) | 8 (13) | 0.51 |
| Non-pulmonary-vein trigger ablation, n (%) | 6 (15) | 16 (26) | 0.19 |

There was no difference in the distribution of ablation lesions during the initial and second ablation between those with and without AF recurrence. In patients with AF recurrence, less frequent use of cryoballoon during the initial ablation and more mapping points during the second ablation were observed. In the second ablation, the majority of patients (73%) had electrical reconnections between left atrium and pulmonary veins, and underwent re-isolation of pulmonary veins.

### Low-voltage areas

ROC analyses demonstrated that a voltage cut-off value of 1.0 mV had a higher area under the curve than that of 0.50 mV (Figure 2). When low voltage was defined as < 1.0 mV, a low-voltage-area size of 5.1 cm^2^ predicted AF recurrence with a sensitivity of 67% and specificity of 64%. Patients with low-voltage areas (< 1.0 mV and ≥ 5 cm^2^) had more frequent AF recurrence (Figure 3 and 4).

**Figure 2.**
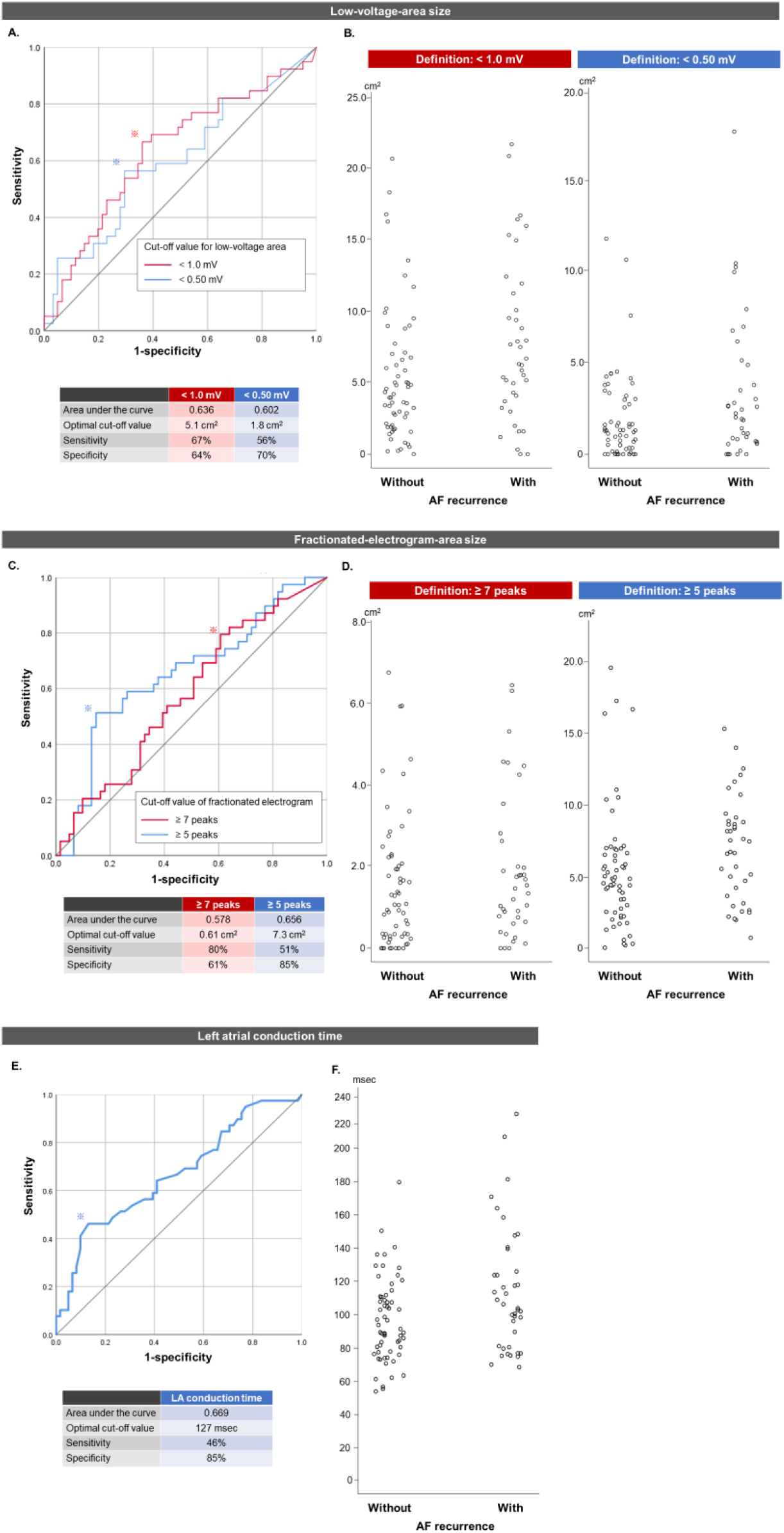
Continuous variables on ROC curves for the prediction of AF recurrence and scatter plots between with and without AF recurrence. Associations between AF recurrence and low-voltage-area sizes using 2 definition of low voltage (A, B), fractionated-electrogram-area size using 2 definitions of number of peaks (C, D), and LA conduction time were depicted on ROC curves and scatter plots. ROC indicates receiver-operator curve.

**Figure 3.**
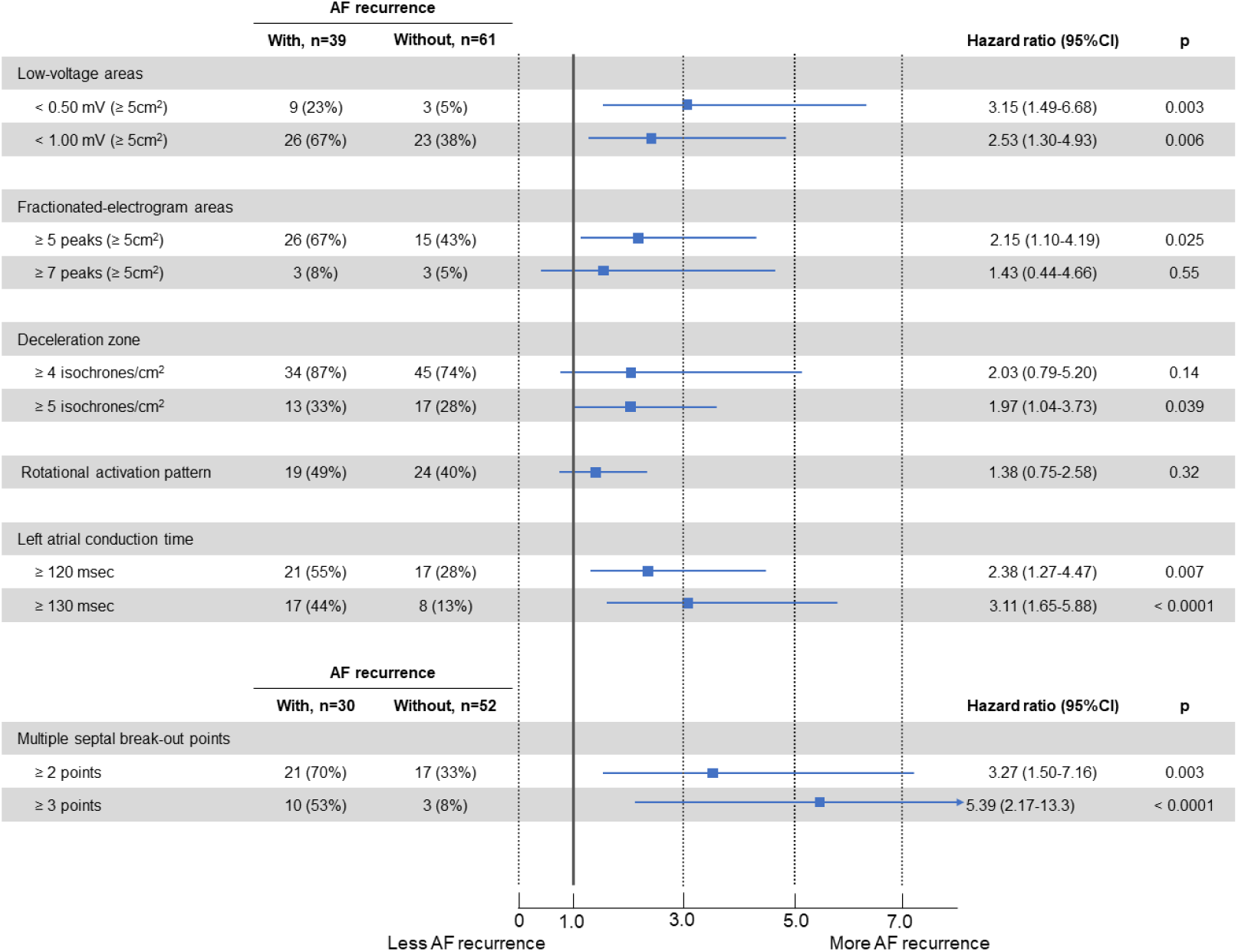
Forrester plots of hazard ratios of abnormal findings on the substrate map for AF recurrence. Hazard ratio, 95% confidence interval, and p value of each parameter for the prediction of AF recurrence are shown. In total study patients, low-voltage areas (< 0.50 mV and < 1.0 mV, ≥ 5 cm^2^), fractionated-electrogram areas (≥ 5 peaks), deceleration zones (≥ 5 isochrones/cm^2^), and prolonged LA conduction time (≥ 120 msec and ≥ 130 msec) were associated with frequent AF recurrence. In patients with substrate mapping under paced beat from the high right atrium, multiple septal break-out points (≥ 2 points and ≥ 3 points) predicted frequent AF recurrence.

**Figure 4.**
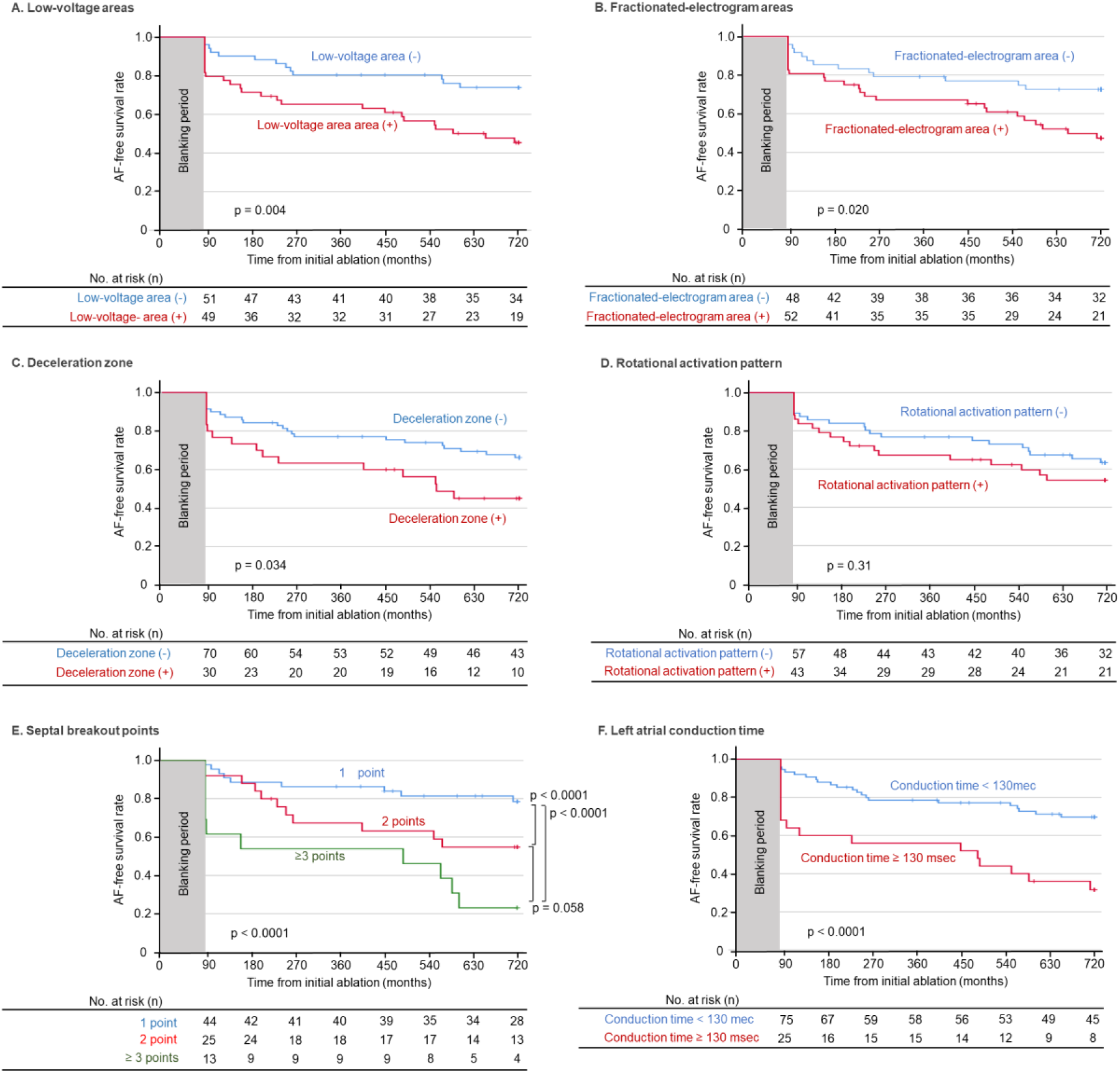
AF-recurrence-free Kaplan-Meier curves stratified by the presence or absence of abnormal findings on the substrate map. Patients with low-voltage areas (< 1.0 mV and ≥ 5 cm^2^), fractionated-electrogram areas (≥ 5 peaks and ≥ 5 cm^2^), deceleration zones (≥ 5 isochrones/cm^2^), multiple septal break-out points (2 points and ≥ 3 points), and prolonged LA conduction time (≥ 130 msec) had lower AF-free survival rates during the 2-year follow-up period. No statistical difference was observed between patients with and without rotation activation pattern.

**Figure 5.**
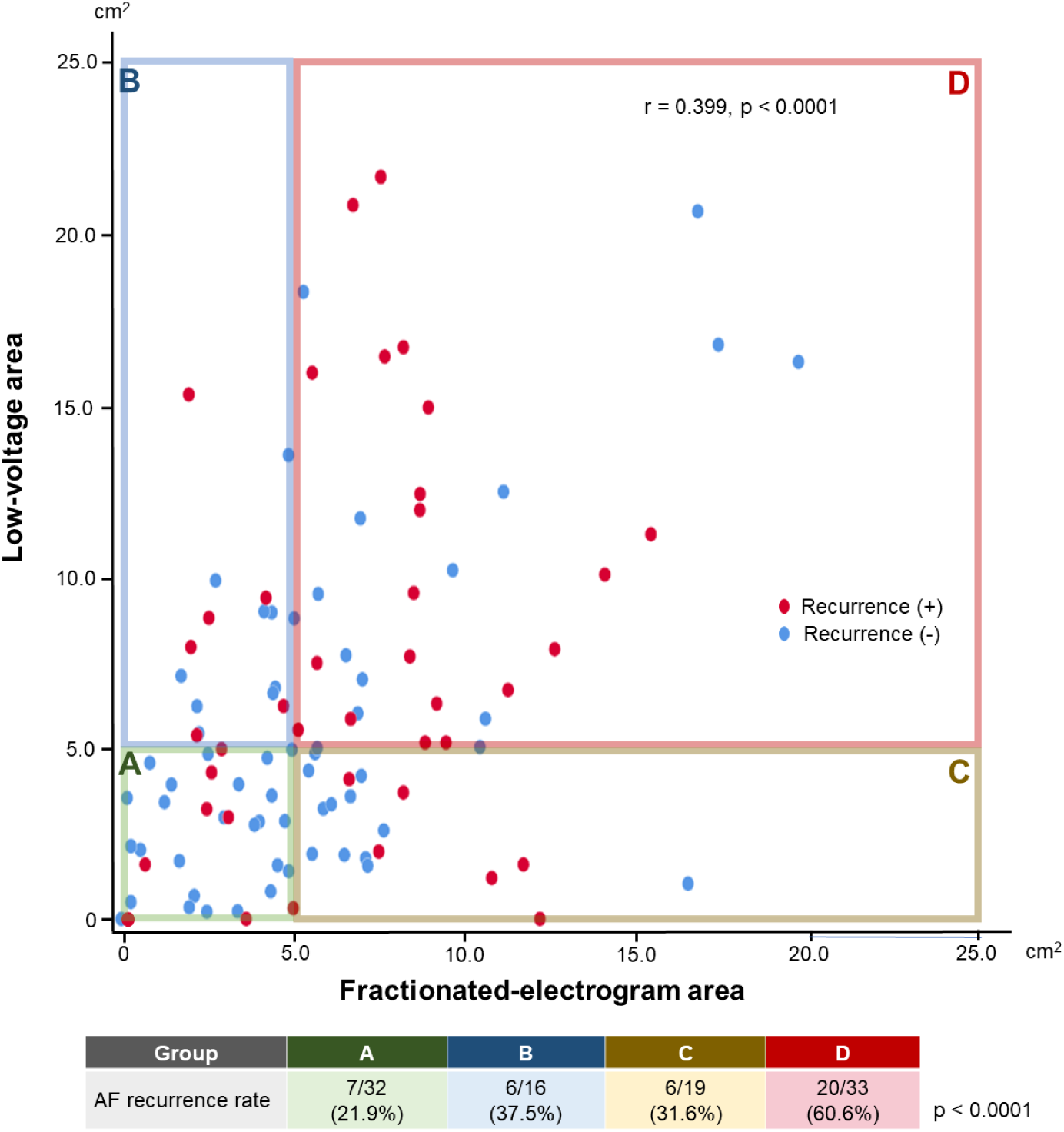
Low-voltage-area size and fractionated-area size in relation to AF recurrence. Scatter plots demonstrate the association between low-voltage-area (< 1.0 mV) size and fractionated-electrogram-area (≥ 5 peaks) size in each patient. Red and blue dots indicate patients with and without AF recurrence. AF recurrence rates were compared between 4 groups (A-D) stratified by the presence of low-voltage area and fractionated-electrogram area. Low-voltage-area and fractionated-electrogram-area sizes showed a weak correlation. Combined presence of the two substrates demonstrated very high AF recurrence rates. AF indicates atrial fibrillation.

### Fractionated-electrogram areas

Fractionated-electrogram areas defined as a peak number of ≥ 5 had a higher area under the curve than those defined as ≥ 7 on ROC analysis for the estimation of AF recurrence (Figure 2), and predicted AF recurrence with a sensitivity of 51% and specificity of 85% when using a cut-off value of 7.3 cm^2^. Patients with fractionated-electrogram areas (≥ 5 peaks and ≥ 5 cm^2^) had more frequent AF recurrence (Figure 3 and 4).

### Deceleration zones

Deceleration zones defined as isochronal crowding of ≥ 4 or ≥ 5 isochrones within a 1-cm radius were observed in 80 (80%) and 30 (30%) patients, respectively. Patients with a deceleration zone (≥ 5 isochrones) has more frequent AF recurrence than those without (Figure 3 and 4).

### Rotational activation pattern

Rotational activation pattern was observed in 43 (43%) patients. By location, rotational activation patterns were predominantly at the anterior region (19 patients) and septal regions (19 patients), followed by the posterior region (6 patients) and roof region (3 patients). There was no statistically significant association between rotational activation pattern and AF recurrence (Figure 3 and 4).

### Multiple septal breakout points

Among 81 patients who underwent mapping under paced beat from the high right atrium, multiple septal break-out points with ≥ 2 points and ≥ 3 points were observed in 38 of 81 (47%) and 13 (16%) patients. Patients with multiple septal break-out points had more frequent AF recurrence than those without (Figure 3 and 4).

### LA conduction time

Patients with AF recurrence had a longer LA conduction time than those without (125 ± 25 vs. 110 ± 18 msec, p = 0.002). ROC analysis revealed that the prediction of AF recurrence using LA conduction time had an area under the curve of 0.669 with an optimal cut-off value of LA conduction time of 127 msec, which predicted AF recurrence with a sensitivity of 46% and specificity of 85% (Figure 2). Patients with LA conduction times of ≥ 130 msec had more frequent AF recurrence than those without (Figure 3 and 4).

### Accumulation of risk factors and AF recurrence

Figure 5 depicts the distribution of low-voltage-area size and fractionated-electrogram-area size in each patient. Low-voltage-area and fractionated-electrogram-area sizes showed a weak correlation (r=0.399, p < 0.001). AF recurrence rates increased in the order of patients without low-voltage areas or fractionated-electrogram areas, those with either, and those with both.

AF recurrence rates stratified by the number of risk factors on a substrate map are shown in Figure 6. Accumulation of these risk factors increased AF recurrence rate in step with a hazard ratio of 1.90, 95% confidence interval of 1.44 - 2.52, p < 0.00001, for each additional risk factor.

**Figure 6.**
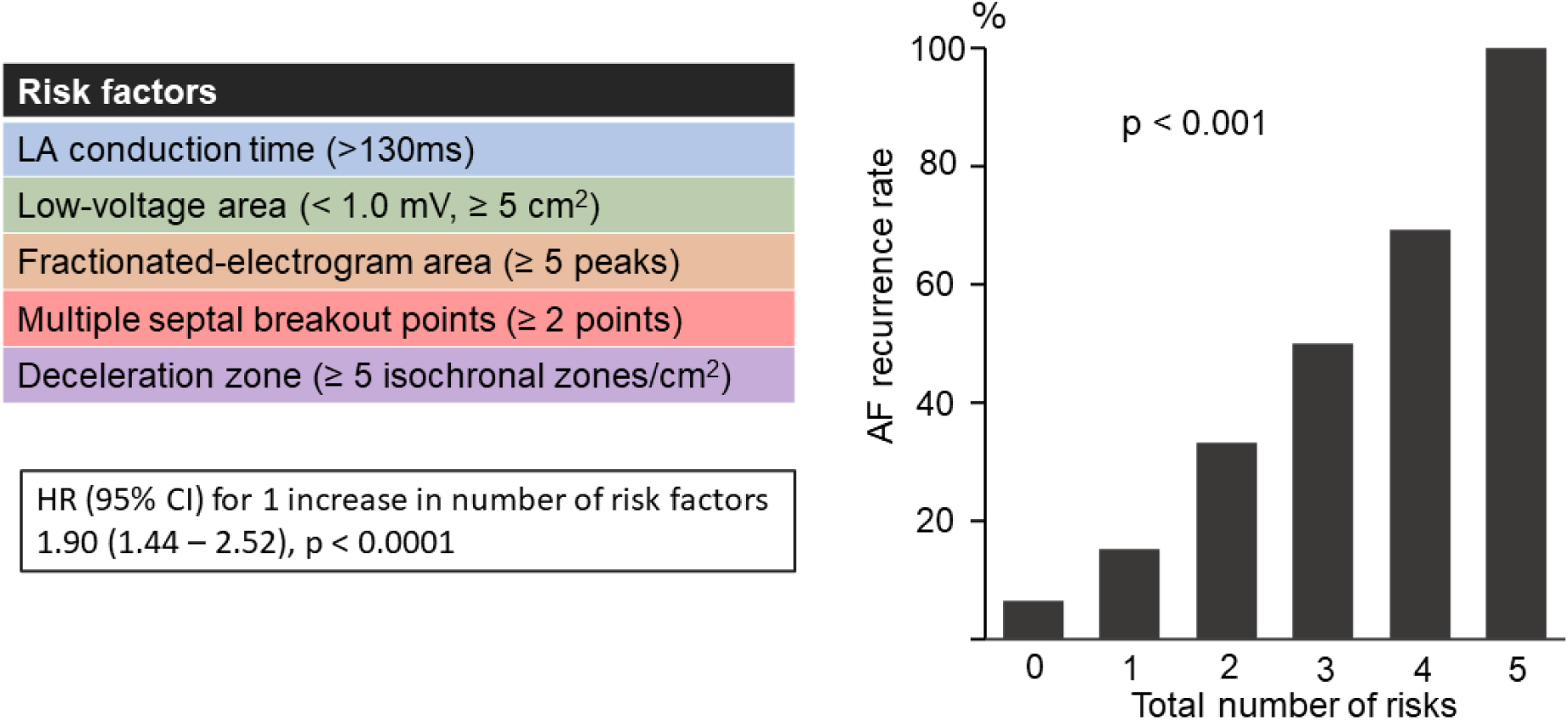
Accumulation of risk factors on the substrate map and AF recurrence. AF recurrence rates stratified by the number of risk factors on the substrate map are shown. Accumulation of these risk factors increased AF recurrence rates in a stepwise manner with an HR = 1.90, 95% CI = 1.44 - 2.52, p<0.00001, for 1 risk increase. AF indicates atrial fibrillation; HR, hazard ratio; CI, confidence interval.

## Discussion

This observational study investigated the impact of LA functional substrates obtained with a high-resolution substrate map on AF recurrence in 100 patients who underwent second AF ablation including pulmonary vein isolation without any additional LA substrate ablation. These patients are thought to have durably isolated pulmonary veins and ablation-naive atrial muscle, which is thought to be ideal for the assessment of atrial arrhythmogenic substrate. The main findings were as follows. Factors associated with 2-year AF recurrence were the presence of low-voltage areas, fractionated-electrogram areas, prolonged LA conduction time, deceleration zone, and multiple septal break-out points. Duplication of these risk factors increased AF recurrence in a step-wise fashion. This is the first study to conclusively assess the prognostic impact of functional substrate of high-resolution maps on AF recurrence after pulmonary vein isolation.

### Low-voltage areas

The presence of low-voltage areas was associated with atrial myocardial fibrotic degeneration based on histopathological analysis of atrial myocardial specimens.^2^ Atrial fibrotic degeneration leads to conduction disturbance and shortening of action potential duration, predisposing to non-pulmonary-vein AF trigger and maintenance of AF reentrant activity.^6–9^

The cut-off value of 1.0 mV for the definition of low-voltage areas had better predictive accuracy than that of the classically used 0.5 mV.^3–5^ This difference is likely attributable to the smaller electrode size of the Orion mapping catheter (printed electrode with 0.4-mm^2^ surface area) than those of the mapping catheters used in most previous studies (≥ 1.0-mm length). The very small printed electrode on the Orion catheter can record higher signal voltage than relatively large electrodes, especially at diseased areas.^10,11^

### Fractionated-electrogram areas

Fractionated electrograms can be generated by diseased myocardium with multiple wave-front propagation and/or conduction disturbance, and are possibly associated with AF development.^12,13^ Precise recording of fractionated electrograms can be achieved by the very small electrode on the Orion catheter with minimal influence of far-field signals and the excellent noise filtering installed on the Rhythmia system. Nevertheless, one problem with utilizing fractionated electrograms as arrhythmogenic substrate is that electrograms with multiple peaks can also be recorded by physiological phenomena such as collision of the propagation wave front and contamination of recording signals due to far-field electrical activities. Selecting a cut-off number of peaks to define the fractionated-electrogram area may help differentiate arrhythmogenic substrate from physiological phenomena. In this study, ≥ 5 peaks of better predicted AF recurrence than ≥ 7. As presented in Figure 1, the fractionated-electrogram area defined using a cut-off value of ≥ 7 was confined, even in the majority of patients with AF recurrence, suggesting that it was too specific to efficiently predict AF recurrence.

A previous study reported that a low-voltage area and fractionated-electrogram area do not always coexist, and sometimes demonstrate different distributions.^14^ The higher AF recurrence rate we saw in patients with both low-voltage areas and fractionated-electrogram areas than in those who had either of them suggests that reduced voltage and electrogram fractionation represent different pathophysiological phenomena, and that their duplication represents an advanced arrhythmogenic substrate.

### Local conduction disturbance

The presence of deceleration zones and multiple septal break-out points, both of which represent local conduction disturbance, was associated with frequent AF recurrence. Local conduction disturbance could not only serve as a slow conduction channel of reentrant regular atrial tachycardia, but could also be associated with LA global substrate predisposing to AF development.

A deceleration zone or localized conduction slowing area first attracted interest in the field of ventricular tachycardia, wherein these findings were associated with a critical isthmus of macro-reentrant ventricular tachycardia.^15,16^ In paroxysmal AF patients, the presence of a deceleration zone was reported to be associated with frequent AF recurrence after pulmonary vein isolation.^17^ Multiple septal breakout points would derive from conduction disturbance of the connection between the right and left atrium, including the Bachman bundle.

In contrast, a rotational activation pattern was not statistically associated with AF recurrence. The somewhat arbitrary judgement of rotational activation might limit the predictive accuracy of this finding.

### Prolonged total LA conduction time

Total LA conduction time is determined by LA conduction speed and distance, both of which are influenced by the severity of myocardial remodeling across the whole LA chamber. Accordingly, prolongation of total LA conduction time would indicate an arrhythmogenic substrate which predisposes to reentrant tachyarrhythmia, including AF. Previous studies have also reported an association between AF recurrence and total atrial conduction times as measured by a 3-dimensional mapping system and P wave duration on the surface electrocardiogram.^18,19^

### Prediction of AF recurrence using functional substrate

In this study, the atrial functional substrate, such as fractionated-electrogram area, total LA conduction time, deceleration zone, and multiple septal break-out points, were individually associated with AF recurrence in patients who underwent pulmonary vein isolation alone. In addition, accumulation of these risk factors increased the risk of AF recurrence in a stepwise manner, possibly representing different electrophysiological phenomena which predispose to AF. Implementation of functional substrate detected by high-resolution mapping enables the identification of patients in whom PVI alone is not enough.

The results of STAR-AF II showed that pulmonary vein isolation without additional substrate ablation is generally appropriate for the catheter ablation of persistent AF.^20^ However, in the clinical setting, pulmonary vein isolation alone does not always achieve sinus rhythm maintenance. Assessment of functional substrate obtained by high-resolution mapping would help identify AF patients who require additional ablation beyond pulmonary vein isolation.

Nevertheless, an important feature of this study is that it did not identify which kinds of outside-of-pulmonary-vein substrate would be ablation targets. Although the possible efficacy of ablation targeting fractionated-electrogram areas or deceleration zones has been reported in single-center clinical studies with small sample sizes, the clinical impact of ablation targeting functional substrate on AF recurrence remains to be determined. Identifying optimal ablation strategies tailored by atrial substrates will require large-scale multicenter randomized controlled trials.

### Limitations

Several limitations of this study warrant mention. First, AF and electrical cardioversion prior to the ablation procedure can affect the LA substrate map. Second, isolation areas encircled by PVI lines were dependent on operator preference, patient anatomy, and ablation device. A wider PVI might include areas with LA arrhythmogenic substrate. Third, AF recurrence after discharge was quantified on the basis of the patient symptoms, giving rise to the possibility that asymptomatic episodes of AF might have been missed. Fourth, the present results are based on a relatively small sample size, retrospective design, and patient enrollment from a single center. Multicenter, prospective, and larger-scale future studies are required to confirm the results.

## Conclusion

In addition to low-voltage areas, we also found that fractionated-electrogram area, prolonged total LA conduction time, deceleration zone, and multiple septal break-out points were associated with AF recurrence after PVI in AF patients. Implementing functional substrate detected by high-resolution mapping enables the identification of patients in whom PVI alone is not enough.

## Data Availability

Data will be available on request to the corresponding author.

---

**Supplementary Figure 1. Patient flow chart**

Among 312 consecutive patients who underwent second ablation for AF, 42 patients examined with another 3-dimensional mapping system, 156 patients with LA ablation outside of the pulmonary vein, and 12 patients without an appropriate substrate map were excluded. The remaining 100 patients were included in the study, and divided into 2 groups: patient with AF recurrence (n=39) and those without (n=61). AF indicates atrial fibrillation; PV, pulmonary vein.

**Supplementary Video 1. Abnormal functional substrate**

A propagation map with multiple septal break-out points and rotational activation pattern is shown.

